# An epigenetic proxy of chronic inflammation outperforms serum levels as a biomarker of brain ageing

**DOI:** 10.1101/2020.10.08.20205245

**Authors:** Eleanor L.S. Conole, Anna J. Stevenson, Claire Green, Sarah E. Harris, Susana Muñoz Maniega, María del. C Valdés-Hernández, Mathew A. Harris, Mark E. Bastin, Joanna M. Wardlaw, Ian J. Deary, Veronique E. Miron, Heather C. Whalley, Riccardo E. Marioni, Simon R. Cox

## Abstract

Low-level chronic inflammation increases with age and is associated with cognitive decline. DNA methylation (DNAm) levels may provide more stable reflections of cumulative inflammatory burden than traditional serum approaches. Using structural and diffusion MRI data from 521 individuals aged 73, we demonstrate that a DNAm proxy of C-Reactive Protein (CRP) shows significantly (on average 6.4-fold) stronger associations with brain structural outcomes than serum CRP. We additionally find that DNAm CRP has an inverse association with global and domain-specific (speed, visuospatial and memory) cognitive functioning, and that brain structure partially mediates this CRP-cognitive association (up to 29.4%), dependent on lifestyle and health factors. These data support the hypothesis that chronic systemic inflammation may contribute to neurodegenerative brain changes which underlie differences in cognitive ability in later life. DNA methylation-based predictors could be used as proxies for chronic inflammatory status.

## Main text

Low-level, systemic chronic inflammation has emerged as a hallmark and potential driver for individual differences in brain ageing, with numerous animal and human studies showing links between sub-acute chronic inflammation and poor brain health^1–7^. While acute inflammation is a healthy, short-term reaction to tissue damage or infection, low-level chronic inflammation is characterised by an ongoing, heightened activity of the immune system that damages cells and tissues. Incidence of peripheral chronic inflammation appears to increase as we age and is strongly associated with age-related diseases including dementia and cardiovascular disease^8^. A striking feature of cognitive ageing at the population level is its wide heterogeneity, with some individuals experiencing more rapid or severe cognitive decline than others^9^ (Figure 1); the determinants of such differences in age-related cognitive decline are not fully understood, but the shift to a progressive, chronic inflammatory state in older-age – termed ‘inflammaging’^10^ – may be a key contributor to inter-individual differences in cognitive ageing.

**Fig. 1.**
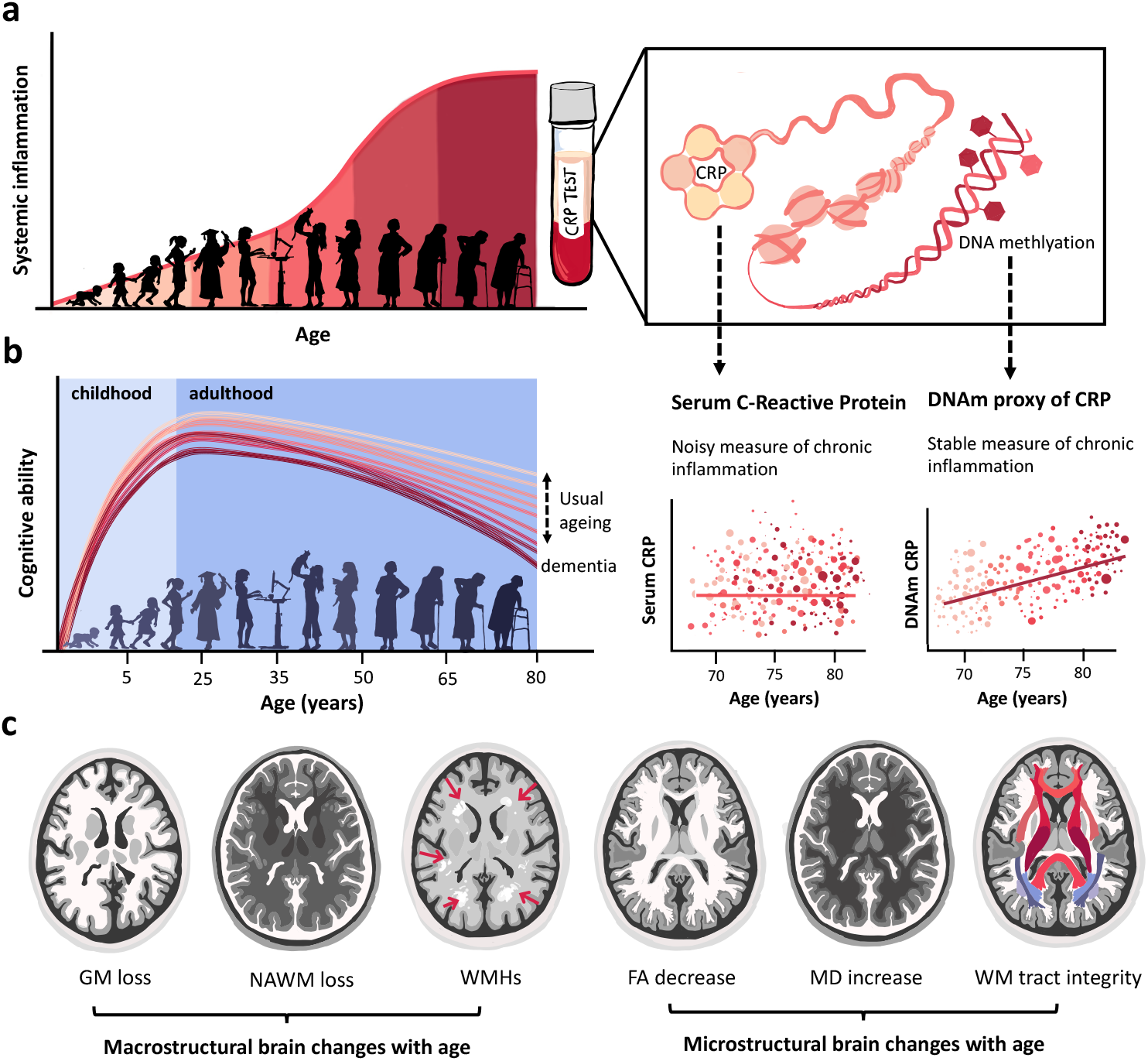
Chronic inflammation increases with age and may contribute to variance in cognitive ability and brain structure. (**a**) Schematic demonstrating how chronic inflammation increases with age (left panel) and can be indexed by inflammatory proteins taken from a blood sample (right panel), such as serum levels and DNA methylation (DNAm) proxies of C-Reactive Protein. Graphs below reflect trajectories of respective inflammation scores over age, as outlined in^21^. (**b**) Life-span curves for cognitive ability, demonstrating that there is considerable inter-individual heterogeneity in rate and timing of cognitive decline, with some people on more accelerated cognitive ageing trajectories than others^9^. (**c**) From left to right, schematic diagram displays structural and diffusion MRI (T1, T2, T2-FLAIR weighted, DTI) correlates of cognitive decline which include alterations in brain macrostructure, such as atrophy in grey matter (GM), normal appearing white matter (NAWM) and increased presence of white matter hyperintensities (WMH; indicated by red arrows). Diffusion tensor imaging (DTI) can reveal alterations in measures of brain microstructure, such as global changes in fractional anisotropy (FA) and mean diffusivity (MD); loss of individual white matter tract integrity can be inferred from probabilistic neighbourhood tractography (PNT) to extract white matter (WM) tracts of interest from DTI data.

However, while chronic inflammation has been independently linked to various neurodegenerative phenotypes^11–14^, the relationship between low level systemic chronic inflammation and cognitive decline in healthy ageing (i.e. in the absence of neurodegenerative disease) is less clearly defined. Studies using serum inflammatory measures in non-clinical groups show mixed positive, negative and null associations with respect to cognitive outcomes^2,15–21^, and have not yet clarified the magnitude and regional extent of brain structural associations^3–6,22,23^. Limitations of previous studies include disparity in methodology (different imaging metrics and cognitive measures), small sample sizes (typically, n <100, although a recent study looked at chronic inflammation in association with cognitive and neuroimaging measures in over 2,000 participants^23^) and a lack of screening for acutely elevated inflammation levels suggestive of infection^24^. Variation in participant health is a likely source of such heterogeneity between study cohorts, as different lifestyle factors are known to increase susceptibility to chronic inflammation (e.g. smoking, obesity, and alcohol consumption) and the related health consequences of inflammation-driven damage (including metabolic syndrome, type-2 diabetes, cardiovascular disease and neurodegenerative pathology) may serve to perpetuate a chronic inflammatory state^8,25^. Moreover, while many studies have investigated the relationship between chronic inflammation and brain health outcomes, few consider all three variables (inflammation, cognitive ability, neuroimaging) concurrently^4^.

Perhaps the largest shortcoming of previous work, however, is how low-level chronic inflammation is measured. As there are no standard biomarkers for low-level chronic inflammation, to date many studies have relied upon canonical blood biomarkers of acute inflammation such as C-Reactive Protein (CRP) to index chronic inflammatory states. A caveat of this methodology is inferring baseline inflammation levels from highly phasic protein levels, which are subject to swift and rapid concentration changes in blood plasma. Serum CRP levels, for example, can increase up to 1,000-fold in response to infection and vary from 1 µg/mL to 500 µg/mL within 24-72h of tissue injury^26^. Thus, the properties which make CRP a good measure of acute infection also introduce significant noise when looking at chronic inflammation associations at the epidemiological level (see Figure 1a). Few studies take repeated measures of serum CRP or attempt to control for this variation when investigating relationships with CRP and brain health phenotypes; where this is collected, longitudinal stability in CRP profiles is low, with large intra-subject variability^27^.

Recent advances in the use of epigenetics to investigate long-term health outcomes^21,28,29^ may present a solution to the challenge of reliably recording average inflammation levels. DNA methylation (DNAm) is an epigenetic mechanism where the addition of a methyl group to a cytosine-phosphate-guanine (CpG) nucleotide base pairing regulates gene activity^30^. DNA methylation (DNAm) profiles are increasingly recognised as biomarkers of disease status and historical archives of environmental exposures^31–33^ and differential DNAm profiles have been identified in inflammatory diseases^34,35^ and inflammation-related disease outcomes^36^. In contrast to genetic variation, DNAm levels vary across the lifecourse^37^ and can be affected by both genes and the environment^38^; measuring DNAm profiles may therefore allow us to tease apart which aspects of lifestyle contribute to chronic inflammation. In addition, while DNAm levels are dynamic, their short term variability is relatively stable^39^, and they may reflect longer-term immune function^40^. In the same cohort as in the present study, a DNAm proxy of CRP exhibited greater longitudinal stability and stronger associations with a single global measure of cognitive functioning than blood based CRP levels^21,41^. Given this temporal stability, a DNAm signature of inflammation could represent a cumulative, aggregate measure of chronic inflammation analogous to the HbA1c test used to index average blood sugar levels for diabetics^21^. We tested this suspected increase in signal:noise conferred by DNAm vs serum CRP on a large number of global and regional measures of brain grey and white matter health to interrogate how inflammation might contribute to cognitive ageing differences.

To date no study has investigated a DNAm measure of CRP relation to structural brain health outcomes. Here we present a detailed comparison of association magnitudes between a serum and DNAm measure of CRP in relation to multiple global and regional structural brain imaging measures, and to various aspects of cognitive function and lifestyle in a well-characterised community-dwelling sample of healthy adults in older age. We hypothesised that our epigenetic score of CRP, created from an out of sample epigenome-wide association study (EWAS), would show significantly stronger associations with brain health than serum levels across a range of detailed neuroimaging and cognitive measures. We additionally investigate whether the association of inflammation with cognitive ability is mediated via alterations to brain structure and how lifestyle factors affect this relationship.

## Results

### DNAm CRP is associated with global and regional brain volume

We studied 521 older adults (aged ∼ 73 years; see Supplementary Table 1) and looked at epigenetic vs serum inflammation associations across a range of cognitive, neuroimaging and lifestyle measures (Supplementary Table 2). To index chronic inflammation, an epigenetic measure of CRP (DNAm CRP) was assembled for each participant: methylation beta values for 7 CpG sites shown to have the strongest association with serum CRP levels were derived, multiplied by their standardised regression weights (taken from the largest meta-EWAS of CRP to date^34^, where CpGs were replicated across a range of cohorts: European, n =8,863, and African-American, n=4,111), and summed to generate a single DNAm CRP score per subject (CpG weights presented in Supplementary Table 5). We found that higher inflammatory burden, indexed by DNAm CRP scores, associated with poor cognitive and neuroimaging brain health outcomes (Supplementary Table 2). DNAm CRP exhibited significantly larger associations with brain structural MRI metrics (including global grey and white matter atrophy, poorer white matter microstructure and increased white matter hyperintensity burden) than serum CRP associations.

Figure 2A illustrates these effect size differences for global neuroimaging measures, which were larger by 6.4-fold, on average, for DNAm CRP vs serum CRP associations. These DNAm CRP-associated brain structural changes were independent of anti-inflammatory drug-use and vascular risk factors (Supplementary Table 5). Participants with a higher inflammatory burden on average had greater overall brain atrophy, with higher DNAm CRP associating with lower total brain volume (β = −0.197, p_FDR_ = 8.42 x 10^−6^), grey matter volume (β = −0.200, p_FDR_ = 1.66 x 10^−5^) and white matter volume (β = −0.150, p_FDR_ = 0.001).

**Fig. 2.**
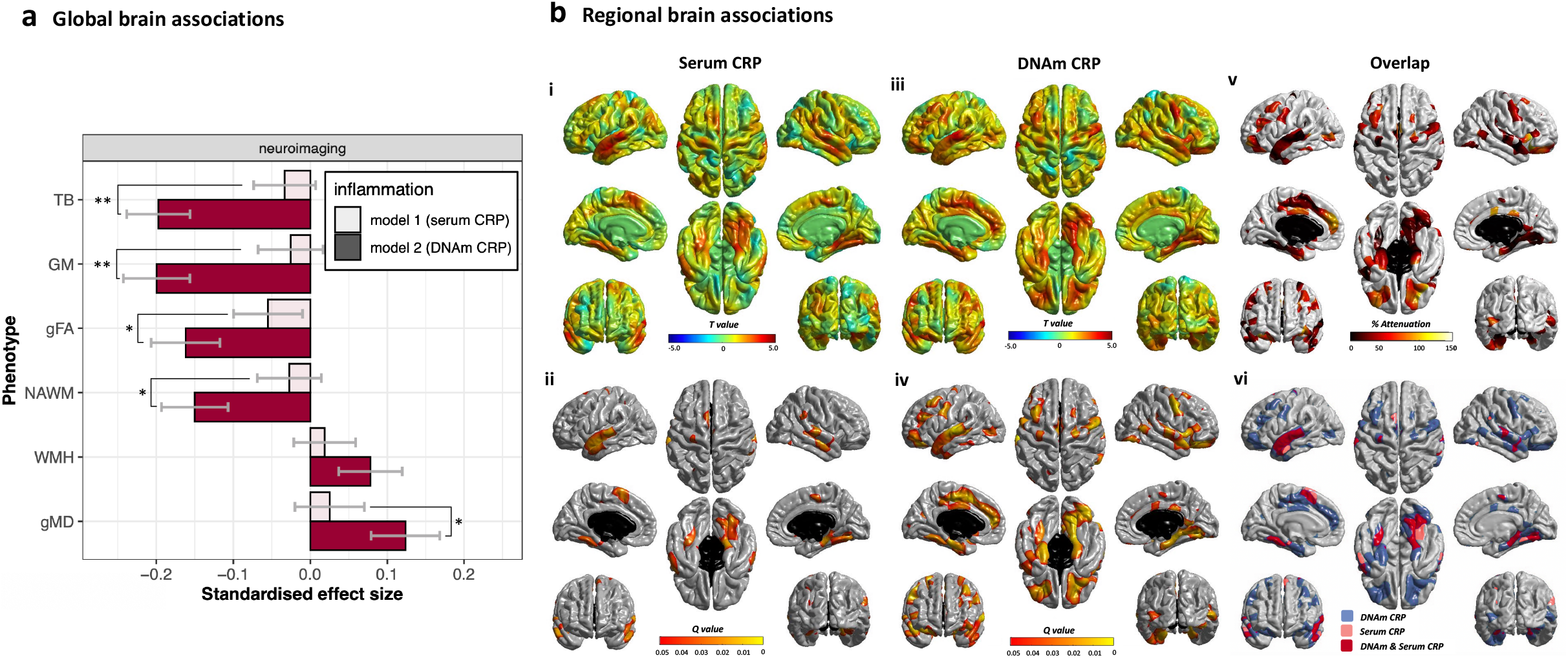
DNAm CRP shows stronger and more widespread associations with global and regional brain structure than serum CRP. (**a**) Associations between CRP measures and brain structure (n = 521); bars show standardised regression coefficients, error bars show standard errors. Asterisks indicate significant differences (*P<0.05, **P<0.01; williams’ test) between serum CRP and DNAm CRP coefficients, which were 6.4-fold larger, on average, for DNAm CRP. (**b**) Regional cortical volume regressed against serum CRP (**i-ii**) and DNAm CRP (**iii-iv**) n = 521. Colours denote the magnitude (T-maps; top) and significance (Q values; bottom) of the negative associations between inflammation and brain cortical volume. Panel (**v**) shows the percentage attenuation for the significant associations between DNAm-CRP and cortical volume when also controlling for serum CRP. Conjunction plot (**vi**) shows the spatial extent of independent contributions and overlap (red) in cortical loci that exhibit FDR-corrected unique associations with simultaneously-modelled serum (pink) and epigenetic (blue) inflammation measures; results are corrected for sex, age and ICV. TB: total brain, GM: grey matter, NAWM: normal-appearing white matter, WMH: white matter hyperintensity, g_f_, general cognitive ability; gFA: general fractional anisotropy, gMD: general mean diffusivity.

After examining global brain structural alterations, we looked at specific regional cortical brain associations with higher inflammation levels. We found regional heterogeneity in the patterning of associations between CRP measures and cortical metrics: atrophy in frontal, anterior lateral and medial temporal lobes were associated with higher DNAm CRP (Figure 2B); inflammation associations with brain cortical thickness are presented in the supplementary document (Supplementary Figure 2). Overall, these results emphasise that the DNAm-CRP score associates with lower cortical volume of specific brain regions (lateral and medial temporal regions of the brain), which show overlap with those of serum CRP and unique variance (Figure 2B-vi), with DNAm CRP reflecting atrophy above and beyond the serum CRP score.

### DNAm CRP is associated with white matter microstructure in specific white matter tracts

Next, we investigated whether higher DNAm CRP was related to lower white matter microstructure based on global and regional diffusion MRI (dMRI) measures by looking at inflammation associations with white matter tract fractional anisotropy (FA; the directional coherence of water molecule diffusion) and mean diffusivity (MD; the magnitude of water molecule diffusion). While serum CRP-dMRI associations were null in all cases (all pFDR > 0.089) (Supplementary Tables 7 and 8), higher DNAm CRP score predicted overall lower general fractional anisotropy (gFA) (β = −0.162, p_FDR_ = 6.94 x 10^−4^) and higher general mean diffusivity (gMD) (β = 0.124, p_FDR_ = 0.010). For specific white matter tracts, the strongest associations were seen for the arcuate fasciculus and uncinate fasciculus, with lower FA and higher MD with higher DNAm CRP (see Figure 3; Supplementary Tables 7-8). For both global and regional measures across grey and white matter, accounting for anti-inflammatory drug status and health and lifestyle covariates did not substantially alter the magnitude or significance of these associations (Supplementary Tables 9 and 10).

**Fig. 3.**
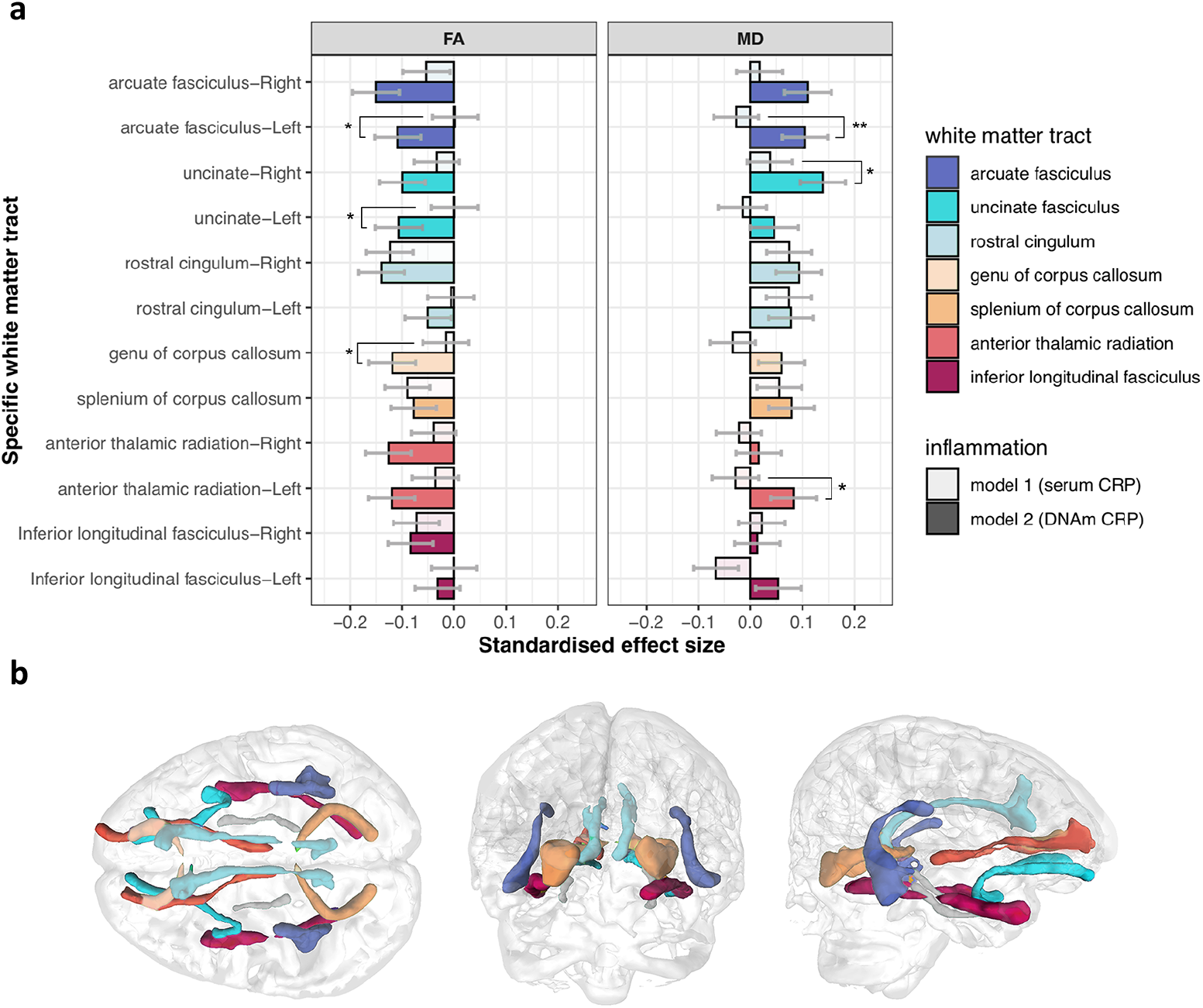
DNAm CRP is associated with white matter microstructure in specific white matter tracts. **(a)** Standardised regression coefficients for associations between white matter tract-averaged fractional anisotropy (FA; left), and mean diffusivity (MD; right). Bars show standardised coefficients and standard errors. Asterisks indicate where associations are significantly larger for DNAm than for serum (*P<0.05, **P<0.01; williams’ test). (arcuate fasciculus n =513; anterior thalamic radiation n = 516; rostral cingulum n=507, genu of corpus callosum n =497; splenium of corpus callosum n =509; inferior longitudinal fasciculus n =516) (**b**) illustration of the respective white matter tracts measured using probabilistic neighbourhood tractography in one LBC1936 study participant.

### Brain structure partly mediates the association of DNAm CRP with cognitive ability

As higher DNAm CRP levels were associated with lower cognitive performance both here (Supplementary Table 2) and previously^21^, we quantified the degree to which brain structural differences contribute to the inflammation-cognition association, and which facets show the strongest unique contributions to this relationship. We used a structural equation modelling (SEM) framework to simultaneously characterise the associations among CRP, brain and cognitive metrics, and also specifically test the hypothesis that brain structure partially and significantly mediates associations between measures of CRP and cognitive ability. Bivariate associations between all variables (inflammation, brain structure, cognitive ability and lifestyle measures) are provided in the Supplementary document (Supplementary Table 8). While total brain (TB) volume, grey matter (GM) volume, normal appearing white matter (NAWM) volume and white matter hyperintensity (WMH) volume all emerged as significant mediators in single SEM models (percentage attenuation 10-21%; Supplementary Table 12), multiple mediator models were used to test the degree to which each global MRI metric contributed uniquely to mediation of the same association (Figure 4d). Here, the sum total of MRI measures significantly mediated the association between DNAm CRP and general cognitive ability (β = −0.047, pFDR = 0.002; percentage attenuation 29.4%). The unique contributions to this variance (Figure 4d and Supplementary Table 13) were largest for NAWM volume (β = −0.029, pFDR = 0.012) indicating that the loss of white matter integrity in particular may contribute to inflammation-associated differences in cognitive functioning in older age. Finally, with the addition of lifestyle and health covariates to our models, no aspect of brain structure remained a significant mediator of the associations between DNAm CRP and general cognitive ability (see Figure 4, Supplementary Tables 12 and 13).

**Fig. 4.**
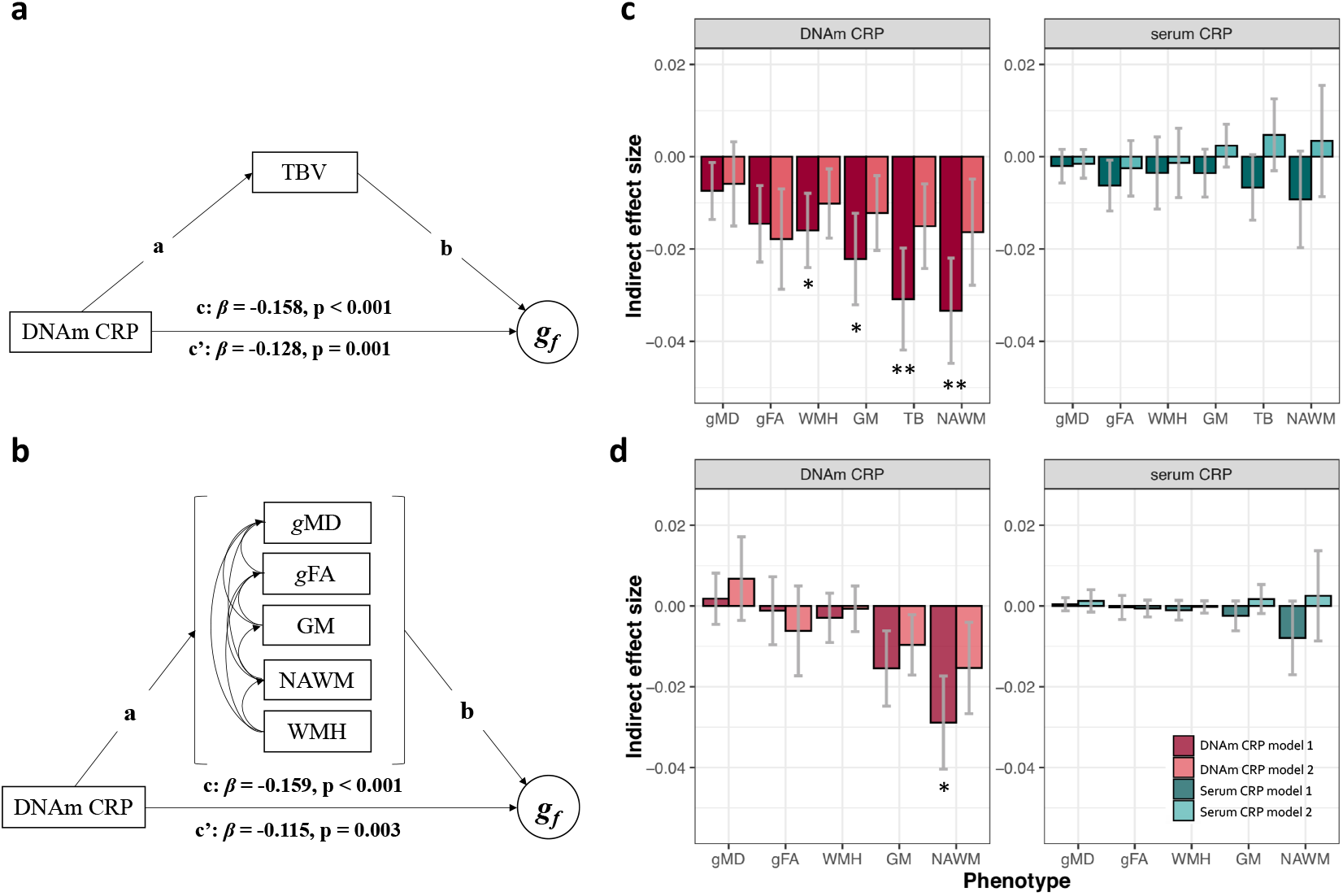
Brain structure partly mediates the association of DNAm CRP with cognitive ability. Top panel (**a-c**) displays single mediator models, bottom panel (**b-d**) displays multiple mediator models. (**a**) Model 1 structural equation model path diagrams showing that that in model 1 the association between DNAm CRP and general cognitive ability (path c) was significantly partially mediated by total brain volume (path ab = −0.031, p = 0.005), attenuating the c path by 19.5% (path c’), and (**b**) 29.4% by multiple MRI variables (ab = −0.047, p = 0.002) (**c**) Single mediator models indirect effect size and standard error bars. (**d**) Multiple mediator models indirect effect size and standard error bars. Light bars show model 1 (includes covariates age and sex), dark bars show model 2 which contains additional health covariates (age + sex + BMI + hypertension + smoking status + alcohol use + CVD history + diabetes). Asterisks denotes FDR p <0.05. TB: total brain, GM: grey matter, NAWM: normal-appearing white matter, WMH: white matter hyperintensity, *g*f, general cognitive ability; gFA: general fractional anisotropy, gMD: general mean diffusivity; n = 521.

## Discussion

In summary, this is the first time that an epigenetic score of CRP has been shown to associate with differences in structural brain measures. We discovered that DNAm CRP shows consistently stronger associations with brain structure than serum CRP (on average, 6.4 fold greater), that these associations are not regionally homogeneous across the brain’s cortex, and that specific aspects of brain structure partly mediate (up to 29.4%) associations between an epigenetic signature of CRP and cognitive functioning, with lifestyle factors significantly attenuating this relationship.

We found regional heterogeneity in the patterning of associations between CRP measures and cortical metrics, indicating differential regional vulnerability to chronic inflammation. Atrophy in frontal, anterior lateral and medial temporal lobes were associated with increased DNAm CRP. The regional overlap between DNAm CRP and serum CRP brain atrophy was strikingly consistent for both cortical thickness and cortical volume, with higher DNAm CRP relating to a thinner cortex and lower volume across a more widespread range of regions, but also identical regions to that of serum CRP. Consistently, previous studies report structural changes associated with inflammatory markers in the temporal and frontal cortices^22,42^. Differential patterns of pro-inflammatory receptor distribution in brain vasculature and tissue are thought to contribute to both local and global brain atrophy^43^, and increased receptor expression may underlie why some brain regions are more vulnerable to inflammation than others. For example, in post-mortem Alzheimer’s disease (AD) patients, pro-inflammatory cytokine receptor density and expression were upregulated in regions of neurodegeneration, including the medial frontal and temporal cortices^44^. Divergent inflammatory markers are also associated with ischemic and haemorrhagic stroke phenotypes, indicating that differing inflammatory pathways and their impact on regional brain vasculature may underlie subtypes of stroke^45^. Although the participants in this study were free of neurodegenerative phenotypes such as stroke or AD, it is possible that sub-incident pathology (undetected lacunar stroke, prodromal dementia) could account for our inflammation-atrophy associations. Moreover, a study on subjects free of history stroke or AD found that CRP was significantly related to progression of carotid atherosclerosis, and that CRP had differential effects in different beds of the arterial brain supply^46^. Overall, it appears that raised levels of inflammatory mediators contribute to localised brain atrophy via their differential and detrimental effects on cerebrovasculature.

Our findings support the hypothesis that chronic systemic inflammation may contribute to changes in brain structure which underlie differences in cognitive ability in later life (see Supplementary Figure 1 for suggested mechanisms). As normal appearing white matter volume emerged as the largest contributor to the mediation of the association between DNAm CRP and a global measure of cognitive ability, we suggest that the brain’s white matter may be particularly vulnerable to the damaging impact of chronic inflammation, and that loss of white matter integrity may drive inflammation-associated accelerated cognitive ageing. Relatedly, several studies have found associations between reduced white matter volume and raised inflammatory mediators both in healthy cohorts^5,19,47^ and those with chronic inflammatory conditions^48^. Possible mechanistic causes for this matter-specific brain atrophy emerge from post-mortem studies looking at microglia deposition: in AD brain samples, microglia activation is specifically increased in white matter relative to grey^49,50^. Chronically elevated inflammation levels in the periphery can cause microglia to shift from a state of comparative quiescence to one of chronic activation^51^, resulting in an enhanced inflammatory response promoting neuronal cell death and subsequent cognitive decline. Moreover, microglia can deteriorate the blood-brain barrier (BBB) from inside the brain, resulting in further infiltration of inflammatory mediators from the systemic circulation (see Supplementary Figure 1)^52^. White matter is supplied by the perforating arterioles and there is evidence that people with greater WMH burden exhibit subtly increased BBB leakage, and in turn, that BBB leakage is associated with worse cognition at a one year follow-up^53^. In a study on patients with systemic inflammatory arthropathies, elevated inflammatory mediators were associated with increased imaging signs of small vessel disease including perivascular spaces and WMH^54^. The areas of brain loss that were particularly associated with the epigenetic CRP here (temporal cortices) are also areas where others have shown increased BBB leakage in persons at risk of AD^53,55^. It therefore seems that damage to the BBB is a possible pathway through which sustained levels of inflammation in the periphery results in neurodegeneration. In addition, increased endothelial inflammation markers have been reported in subjects with increased WMHs^56^, providing more evidence for neurovascular mediated neuroinflammation and subsequent downstream white matter loss.

In line with this, we found that higher DNAm CRP is related to ostensibly poorer white matter microstructure (lower FA and higher MD). In particular, the white matter tracts of arcuate fasciculus and uncinate fasciculus showed the most consistent significant relationships with DNAm CRP levels (across both FA and MD), alongside significantly lower FA in the anterior thalamic radiation, which is susceptible to the effects vascular risk and cognitive ageing^57^. Similar cohorts also show links between white matter integrity and chronic inflammation: a faster decline in serum CRP levels was related to greater white matter tract health, with declines in inflammation over six years predicting FA in various white matter tracts^47^. Similarly, higher midlife CRP levels predicted reduced white matter integrity in later life^5^. As with discrete regional cortical thinning, our results indicate that loss of white matter integrity occurs at both regional and global levels, indicating that individual white matter tracts may have differential vulnerability to the effects of chronic inflammation. Stronger microstructural associations with DNAm CRP than serum CRP point towards potential utility in monitoring chronic inflammation in AD or small vessel disease populations via epigenetic markers to characterise and quantify inflammatory status.

Given that all mediations via brain structure were significantly attenuated by the addition of lifestyle factors into our models, it is clear that various exposures and predisposing health attributes influence the association between chronic inflammation and brain health. These included factors that showed significant associations with higher inflammation levels (BMI, diabetes, smoking, alcohol consumption and hypertension) in this study (see Supplementary Table 2) and others^8,58^. Many of the lifestyle attributes associated with chronic inflammation (and probable perpetuators of it) are at least partly modifiable, yet studies that have successfully targeted these risk factors and shown corresponding reductions in inflammation levels are scarce^8^. Variation in serum levels may be confounding this relationship, and our data suggest that the DNAm-based predictor may act as a quantifiable archive of the longitudinal effects of these exposures, and other unaccounted for health and genetic profiles, that serum CRP levels fail to capture.

The strengths of the present work include the large sample size, array of multi-modal data, narrow age range and ethnic homogeneity of our cohort; the concurrently-collected data were optimal for interrogating the relationship between chronic inflammation and cognition from neuroimaging, cognitive, epigenetic and lifestyle angles. Compared to recent EWAS-neuroimaging research, this study is exceptionally well-powered with 521 participants after exclusions^29^. The narrow age-range and ethnic homogeneity are simultaneously limitations of this study, as they restrict the degree to which our findings can be related to other populations and limit our scope to identify inflammation-brain health associations at other times of life. Equally, given the cross-sectional nature of this study, we are unlikely to capture the effect of more age-related changes in inflammatory profile and cognitive decline. Observed cognitive and brain structural alterations could be independently related to a genetic-predisposition unaccounted for by our health and lifestyle covariates, such as metabolic-syndrome or more niche vascular vulnerability. Although we endeavoured to remove participants with cognition-related pathology (stroke, AD, Parkinson’s disease and MCI), these were screened via self-reported diagnoses and we may be missing undiagnosed or subclinical incident neurodegenerative pathology. While it is exciting to consider that DNAm levels could provide more accurate reflections of chronic inflammatory status, more work is required to determine the direction and strength of this association with brain health phenotypes. It is possible that chronic inflammation is not a cause, rather a marker of, or even a response to, unrelated neurodegenerative phenotypes that can lead to changes in cognitive ability. To disentangle directionality of effects, there is demand for studies that collect repeated measures of DNAm, cognitive and neuroimaging data over time. A few cohorts, including LBC1936, are suited to this type of longitudinal study as highlighted in a recent systematic review^59^; to further strengthen causal inference or in cases where no such data is available, a two-sample Mendelian Randomisation approach may be best suited.

These findings have clinical implications, for example, relating DNAm proxies of inflammatory exposure to MRI patterns in neonates could shed light on the role of chronic inflammation and brain structure in preterm infants, where chronic inflammation is of known brain health consequence but disentangling maternal vs infant exposure is difficult^60^. Future studies should consider examining a wider range of DNAm inflammatory markers (DNAm levels of interleukins, prostaglandins, neurotrophins); looking at DNAm inflammatory markers in younger participants (where there is likely greater variation in baseline inflammation levels); and looking at DNAm inflammatory markers in specific brain pathology cases (e.g. multiple sclerosis patients).

In conclusion, these findings support the hypothesis that chronic systemic inflammation may contribute to neurodegenerative brain changes which underlie differences in cognitive ability in later life. Previous studies exploring this relationship may underestimate the brain and cognitive sequelae of chronic inflammation by relying on single measurements of phasic serum proteins. By utilising an epigenetic inflammation measure, which integrates information from multiple immune-related CpG sites, we may provide a more reliable measure of chronic inflammation and thus a more comprehensive overview of the consequences of chronic inflammation on brain structure and function. Reliable monitoring of inflammatory exposure could enable clinicians to review the efficacy of drug and lifestyle interventions to attenuate inflammation levels with a view to improving cognitive outcomes.

## Materials and Methods

### Participants

Data are drawn from the Lothian Birth Cohort 1936 (LBC1936). The cohort is composed of 1,091 individuals, born in 1936 and re-contacted around 60 years later (mean age: 69.6 ± 0.8 years). Most took part in the Scottish Mental Survey 1947 at age 11. Detailed cognitive, genetic, epigenetic, health and lifestyle data was collected at this first wave, and also three years later, supplemented with a detailed structural brain imaging protocol (see below). Participants were free of neurodegenerative diagnoses at baseline and, as the study’s focus is on healthy cognitive ageing, were excluded if they had a self-reported history of stroke (n=55), Parkinson’s (n= 5), or dementia (n=2) or had an MMSE < 24, indicating mild cognitive impairment (n=7). We additionally excluded participants with serum CRP level >10mg/L (n = 32), suggestive of acute infection or illness at the time of blood draw ^24^. After exclusions, a total of 680 participants had DNAm CRP data at age 73 years, 521 of whom also provided brain MRI data. We used the maximum available sample size in all analyses. All variables described in this study were collected at Wave 2.

### Brain imaging data

Structural and diffusion tensor (DTI) MRI acquisition and processing in LBC1936 were performed according to an open-access protocol ^61^. A 1.5 T GE Signa HDx clinical scanner (General Electric, Milwaukee, WI, USA) was used to collect structural T1-, T2-, T2*-, and fluid attenuated inversion recovery-weighted images. Total brain (TB), grey matter (GM), normal-appearing white matter (NAWM) and white matter hyperintensity (WMH) volumes were segmented using a semi-automated multi-spectral technique ^62^. Local processing and QC of cortical reconstruction and segmentation was performed using FreeSurfer v5.1 on T1-weighted volumes. Following visual inspection of the outputs (to check for aberrant surfaces and tissue segmentation failures, which were removed from analysis) these were registered to the fsaverage surface. White matter connectivity data – measures of fractional anisotropy (FA) and mean diffusivity (MD) – were created and segmented using the BEDPOSTX/ProbTrackX algorithm in FSL (https://fsl.fmrib.ox.ac.uk) and Tractor (https://www.tractor-mri.org.uk). Using probabilistic neighbourhood tractography (PNT), tract-average white matter FA and MD were derived as the average of all voxels contained within the resultant tract maps (genu of corpus callosum; splenium of corpus callosum; arcuate fasciculus; anterior thalamic radiation; rostral cingulum; uncinate fasciculus; inferior longitudinal fasciculus), as described previously ^63,64^. A general factor of FA (g_FA_) and MD (g_MD_) was derived for each participant from the first un-rotated principal component of a principal components analysis (PCA) on twelve of the white matter tracts FA and MD values; participants with up to 2 missing values from specific tracts had data replaced with the mean value for that tract. These general factors reflect common microstructural properties across main white matter pathways and capture the common variance in white matter integrity. Details of individual test loadings are provided in the supplementary document (Supplementary Table 3).

### C-Reactive Protein data

Serum CRP was measured from whole-blood samples using a high sensitivity assay (enzyme-linked immunosorbent assay; R&D Systems, Oxford, UK) ^54^.

### DNA methylation preparation and DNAm CRP score

Genome-wide DNA methylation was measured in blood samples using the Illumina Human MethylationEPIC BeadChip at the Edinburgh Clinical Research Facility Genetics Core; details of this profiling has been outlined^65^. The DNAm CRP score was calculated for each participant as described previously^66^, briefly, a DNAm CRP score was assembled for each participant in Wave 2 of LBC1936; this was created by means of a weighted composite score, based on a discovery meta-analysis (9 cohorts, n =8,863) and a replication meta-analysis (4 cohorts, n = 4111) of CRP-EWAS studies ^34^. Methylation beta values were derived for the 7 CpG sites shown to have the strongest association with serum CRP levels, and then multiplied by their standardised regression weights (taken from the meta-EWAS of CRP;^34^)and added together. Given that all regression weights from the EWAS were negative, a higher DNAm CRP score (i.e closer to 0) corresponds to a higher inflammatory profile. Relative weights for the 7 CpGs are included in the supplementary document (Supplementary Table 5).

### Cognitive ability data

A general fluid-type cognitive ability score (*g*f) was derived from the first un-rotated principal component of a PCA of relevant cognitive tests from the Wechsler Adult Intelligence Scale-Third Edition (WAIS-III^UK^)^67^. Relevant cognitive tests and individual weightings of *g*f and the latent variables of processing speed, visuospatial ability and verbal memory can be found in Supplementary Table 4.

### Lifestyle variables

Lifestyle variables included body mass index (BMI; kg/m^2^) alongside variables relating to self-reported health and disease history: cardiovascular disease history (CVD); hypertension; diabetes; smoking status (coded as current smoker [1] versus ex/non-smoker [0]) and current alcohol use (alcohol units per week).

### Volumetric brain associations with inflammation

Linear regression models were used to identify the proportion of phenotypic variance explained by DNAm CRP and to determine whether this was independent of the serum CRP signal for each neuroimaging, cognitive and lifestyle phenotype. Logistic regressions were conducted for self-reported disease history variables as these had binary outcomes (disease/no disease). The phenotypic measure was the dependent variable, and the serum CRP or DNAm CRP score was the independent variable of interest. Differences between association magnitudes (serum CRP vs epigenetic CRP associations) were assessed using the Williams’ test for dependent groups with overlapping correlations (cocor.indep.groups.overlaps) as implemented in the cocor R package (http://cran.r-project.org/web/packages/cocor/cocor.pdf).

### Regional brain analyses

Localized associations between DNAm CRP score and vertex-wise cortical volume, area and thickness were performed using linear regression, controlling for age, sex, and ICV. We used the SurfStat MATLAB toolbox (http://www.math.mcgill.ca/keith/surfstat) for Matrix Laboratory R2012a (The MathWorks, Inc., Natick, MA, USA). The resulting statistical maps (*t*-maps) were corrected for multiple comparisons using FDR with a q-value of 0.05 across all 327,684 vertices on the cortical surface. Negative associations with CRP measures (e.g. lower volume with higher inflammation) were represented by the hot end of the colour spectrum. Following exclusions based on a history of stroke, dementia or an MMSE < 24, n = 521 had complete vertex-wise MRI, epigenetic and phenotypic data. White matter tract-specific associations with CRP measures were investigated using regression models adjusted for age and sex as described previously for global volumetric measures (TB, GM, NAWM, WMH).

### Specific white matter tract associations with inflammation

As with volumetric brain-inflammation associations, regressions were run with regional FA and MD values of left and right projections of the individual white matter tracts (genu of corpus callosum; splenium of corpus callosum; arcuate fasciculus; anterior thalamic radiation; rostral cingulum; uncinate fasciculus; inferior longitudinal fasciculus) as dependent variables.

### Sensitivity analyses

We sought to investigate whether anti-inflammatory drug status had an influence on our models. In a sensitivity analysis, anti-inflammatory drug status (collected at baseline and coded as a dichotomous variable: on medication = 1; not on medication = 0) was included as a covariates alongside age and sex. Similarly, we included lifestyle and health covariates separately in models (alongside age and sex) to determine whether individual aspects of health and lifestyle had an impact on the association of inflammation with brain-health phenotypes.

### Mediation analyses

We ran mediation analyses in a structural equation modelling (SEM) framework using the R ‘lavaan’ package (https://cran.r-project.org/web/packages/lavaan/lavaan.pdf). This simultaneously characterised associations among CRP, brain and cognitive metrics, and also specifically tested the hypothesis that brain structure would partly and significantly mediate associations between measures of CRP and cognitive ability. Both single and multiple mediator models were specified (see Figure 4a-b as example). Single mediator models provided information on the proportion of CRP-cognitive associations attributable to individual neuroimaging metrics. By contrast, in Multiple mediator models, brain structural variables were entered simultaneously as covarying mediators. This allowed us to quantify the proportion of variance in CRP-cognitive associations uniquely explained by each facet of brain structure (GM, NAWM, WMH, gFA, gMD). The primary estimates of interest in this study are the degree of change (mediation) in the direct path (c to c’) between inflammation measures (DNAm CRP or serum CRP) and cognitive ability when the indirect path from inflammation to cognitive ability via brain structure (a x b) is included. A significant mediation of the c path (to c’) is denoted by the statistical significance of this indirect effect. Bootstrapping was used calculate the standard errors. Multiple comparisons were corrected for by FDR correction. These mediations were re-run when accounting for self-reported health variables as covariates. In Model 1 age and sex were covariates; in Model 2, they were age, sex, BMI, hypertension, diabetes, smoking status and alcohol use. Model fit was evaluated based on root mean squared error approximation (RMSEA), the comparative fit index (CFI), the standardized root mean square residual (SRMR) and the Tucker– Lewis index (TLI). We considered a model an acceptable fit when it respected the following thresholds: RMSEA ≤ 0.05; SRMR ≤ 0.06; CFI ≥ 0.97; TLI ≥ 0.95 as recommended by ^68^.

### Statistical Analyses

Statistical analyses were performed in R version 3.6.1 (https://www.rproject.org). Alpha was 0.05 for all analyses and results were corrected for multiple comparisons using the false discovery rate (FDR) using the ‘p.adjust’ function in the ‘stats’ package in R. Standardized coefficients are reported throughout to facilitate comparison of associations. Serum measures of CRP were log-transformed to correct a positively skewed distribution. When conducting analysis for brain structural associations, to control for the confounding effect of head size, all global MRI volumetric measures (TB, GM, NAWM, WMH) were corrected for intracranial volume (ICV) and expressed as a ratio of ICV. Pairwise bivariate associations were assessed between markers of inflammation, neuroimaging and lifestyle covariates using Pearson correlation.

## Data Availability

The data analysed in this study is not publicly available as it contains data that could compromise participant consent and confidentiality, but can be requested via a data access request to the Lothian Birth Cohorts research group.

## Acknowledgements

We thank the Lothian Birth Cohort 1936 members who took part in this study, and Lothian Birth Cohort 1936 research team members who collected, entered and checked data used in this manuscript. The LBC1936 and this research are supported by Age UK (Disconnected Mind project) and by the UK Medical Research Council [MRC; G0701120, G1001245, MR/M013111/1, MR/K026992/1]. E.L.S.C. and A.J.S. are supported by funding from the Wellcome Trust 4-year PhD in Translational Neuroscience [108890/Z/15/Z to E.L.S.C.; 203771/Z/16/Z to A.J.S]. M.E.B., I.J.D. and S.R.C. are also supported by a National Institutes of Health (NIH) research grant R01AG054628.

## Author contributions

E.L.S.C., S.R.C. and R.E.M. designed the analysis. E.L.S.C. conducted the analysis and drafted the work. A.J.S. prepared the DNA methylation data. S.M.M. provided the illustration in Figure 3B. S.E.H., S.M.M., M.V.H., M.A.H., M.E.B., J.M.W., I.J.D., and S.R.C. collected or analysed the brain MRI and/or cognitive data. All authors critically revised the work and have approved the submitted version. Data Availability

## Materials and correspondence

To be addressed to Dr. Simon Cox

