## Supplementary Material for "An epigenetic proxy of chronic inflammation outperforms serum levels as a biomarker of brain ageing"

**This document includes:**

Supplementary Tables 1-13
Supplementary Figures 1-2

### Supplementary Tables

#### Table 1. Participant characteristics

| Class | Variable | Units | mean | SD | n |
|---|---|---|---|---|---|
| **Demographics** | sex | M: F | 358 : 322 | - | 680 |
| | Age | years | 72.494 | 0.716 | 680 |
| **Inflammation** | DNAm CRP | Std units | -0.018 | 0.001 | 680 |
| | serum CRP (high sensitivity) | mg/L | 2.016 | 1.92 | 680 |
| **Neuroimaging** | TB | $cm^3$ | 992.03 | 89.8 | 521 |
| | GM | $cm^3$ | 473.45 | 45.1 | 521 |
| | NAWM | $cm^3$ | 477 | 50.67 | 521 |
| | WMH | $cm^3$ | 8.80 | 1.31 | 517 |
| | gFA | Std units | 0.04 | 0.97 | 496 |
| | gMD | Std units | -0.023 | 0.97 | 496 |
| **Fractional anisotropy** | Genu of corpus callosum | Std units | 0.376 | 0.046 | 497 |
| | Splenium of corpus callosum | Std units | 0.511 | 0.066 | 509 |
| | Left Arcuate Fasciculus | Std units | 0.436 | 0.039 | 513 |
| | Right Arcuate Fasciculus | Std units | 0.415 | 0.039 | 485 |
| | Left Anterior Thalamic Radiation | Std units | 0.324 | 0.032 | 501 |
| | Right Anterior Thalamic Radiation | Std units | 0.335 | 0.035 | 516 |
| | Left Rostral Cingulum | Std units | 0.437 | 0.058 | 507 |
| | Right Rostral Cingulum | Std units | 0.41 | 0.047 | 500 |
| | Left Uncinate fasciculus | Std units | 0.325 | 0.034 | 485 |
| | Right Uncinate fasciculus | Std units | 0.32 | 0.031 | 506 |
| | Left Inf. Longitudinal Fasciculus | Std units | 0.386 | 0.051 | 516 |
| | Right Inf. Longitudinal Fasciculus | Std units | 0.377 | 0.048 | 515 |
| **Mean diffusivity** | Genu of corpus callosum | Std units | 798.7 | 78.3 | 497 |
| | Splenium of corpus callosum | Std units | 809 | 123.6 | 509 |
| | Left Arcuate Fasciculus | Std units | 658.5 | 50.4 | 513 |
| | Right Arcuate Fasciculus | Std units | 644.8 | 50.6 | 485 |
| | Left Anterior Thalamic Radiation | Std units | 756.5 | 63.9 | 501 |
| | Right Anterior Thalamic Radiation | Std units | 739.1 | 76.2 | 516 |
| | Left Rostral Cingulum | Std units | 636.1 | 45.3 | 507 |
| | Right Rostral Cingulum | Std units | 622.4 | 44.1 | 500 |
| | Left Uncinate fasciculus | Std units | 764.6 | 57.7 | 485 |
| | Right Uncinate fasciculus | Std units | 760.6 | 49 | 506 |
| | Left Inf. Longitudinal Fasciculus | Std units | 772.7 | 105.4 | 516 |
| | Right Inf. Longitudinal Fasciculus | Std units | 757.9 | 97.1 | 515 |
| **Cognitive** | *gf* | Std units | 0.052 | 0.975 | 677 |
| | processing speed | Std units | 0.058 | 0.947 | 661 |
| | visuospatial ability | Std units | 0.048 | 0.997 | 676 |
| | verbal memory | Std units | 0.04 | 0.972 | 667 |
| **Lifestyle** | hypertension | % Yes | 53% | - | 362 |
| | CVD | % Yes | 29% | - | 197 |
| | diabetes | % Yes | 10% | - | 71 |
| | alcohol units per week | Std units | 14.16 | 14.89 | 533 |
| | smokers | % Yes | 8% | - | 55 |
| | ex smokers | % Yes | 44% | - | 301 |
| | BMI | Kg/m2 | 27.769 | 4.252 | 680 |
| | anti-inflammatory medication | % Yes | 7.21% | | 49 |

TB: total brain, GM: grey matter, NAWM: normal-appearing white matter, WMH: white matter hyperintensity, gf, general cognitive ability; gFA: general fractional anisotropy, gMD: general mean diffusivity.

**Table 2. Cross-sectional associations. Serum CRP and epigenetic CRP as predictors of health outcomes.**

| | Phenotype | Serum CRP (model 1) | | | DNAm CRP (model 2) | | | Δ association magnitudes |
|---|---|---|---|---|---|---|---|---|
| | | β | SE | p | β | SE | p | p |
| Neuroimaging | TB | -0.033 | 0.040 | 0.579 | -0.197 | 0.041 | **$8.42 \times 10^{-6}$** | **0.001** |
| | GM | -0.026 | 0.042 | 0.670 | -0.200 | 0.043 | **$1.66 \times 10^{-5}$** | **0.001** |
| | NAWM | -0.027 | 0.042 | 0.657 | -0.150 | 0.043 | **0.001** | **0.015** |
| | WMH | 0.018 | 0.042 | 0.740 | 0.108 | 0.043 | **0.017** | 0.069 |
| | gFA | -0.055 | 0.045 | 0.347 | -0.162 | 0.045 | **$6.97 \times 10^{-4}$** | **0.032** |
| | gMD | 0.025 | 0.045 | 0.677 | 0.124 | 0.045 | **0.010** | **0.049** |
| Cognitive | visuospatial ability | -0.082 | 0.038 | 0.069 | -0.097 | 0.037 | **0.014** | 0.213 |
| | processing speed | -0.088 | 0.038 | 0.054 | -0.144 | 0.038 | **$4.64 \times 10^{-4}$** | 0.267 |
| | verbal memory | -0.046 | 0.038 | 0.347 | -0.095 | 0.038 | **0.017** | 0.175 |
| | gf | -0.098 | 0.038 | **0.027** | -0.158 | 0.037 | **$6.55 \times 10^{-5}$** | 0.732 |
| Lifestyle / health | diabetes | 0.030 | 0.039 | 0.601 | 0.110 | 0.038 | **0.007** | 0.070 |
| | BMI | 0.291 | 0.036 | **$5.88 \times 10^{-14}$** | 0.187 | 0.037 | **$3.06 \times 10^{-6}$** | **0.014** |
| | smoking status | 0.136 | 0.037 | **$9.69 \times 10^{-4}$** | 0.230 | 0.037 | **$1.16 \times 10^{-8}$** | **0.030** |
| | CVD | 0.007 | 0.038 | 0.856 | 0.102 | 0.037 | **0.010** | **0.031** |
| | hypertension | 0.049 | 0.038 | 0.339 | 0.055 | 0.038 | 0.168 | 0.903 |
| | alcohol units per week | 0.058 | 0.042 | 0.311 | 0.206 | 0.040 | **$2.06 \times 10^{-6}$** | **0.003** |

Note: Bold typeface denotes $p < 0.05$. Asterisks indicate significant differences between serum and DNAm regressions. TB: total brain, GM: grey matter, NAWM: normal-appearing white matter, WMH: white matter hyperintensity, gf, general cognitive ability; gFA: general fractional anisotropy, gMD: general mean diffusivity. All p values reported are FDR corrected.

**Table 3: Tract loadings for general factors of white matter fractional anisotropy and mean diffusivity**

| *White matter tract* | *PC1(FA)* | *PC1 (MD)* |
|---|---|---|
| Genu of corpus callosum | 0.647 | 0.646 |
| Splenium of corpus callosum | 0.490 | 0.308 |
| Left arcuate fasciculus | 0.722 | 0.735 |
| Right arcuate fasciculus | 0.69 | 0.767 |
| Left anterior thalamic radiation | 0.65 | 0.72 |
| Right anterior thalamic radiation | 0.634 | 0.645 |
| Left rostral cingulum | 0.55 | 0.648 |
| Right rostral cingulum | 0.573 | 0.752 |
| Left uncinate fasciculus | 0.637 | 0.665 |
| Right uncinate fasciculus | 0.669 | 0.744 |
| Left Inferior longitudinal fasciculus | 0.494 | 0.41 |
| Right Inferior longitudinal fasciculus | 0.477 | 0.371 |
| **Proportion of variance** | **0.370** | **0.405** |

**Table 4: Test loadings for general factors of cognitive ability measures**

| *Cognitive test* | ***gf**** | **processing speed** | **visuospatial ability** | **verbal memory** |
|---|---|---|---|---|
| digit-span backwards | 0.634 | - | - | 0.608 |
| symbol search | 0.746 | 0.816 | - | - |
| digit-symbol coding | 0.739 | 0.834 | - | - |
| matrix reasoning | 0.691 | - | 0.796 | - |
| letter-number sequencing | 0.708 | - | - | - |
| block design | 0.71 | - | 0.839 | - |
| Inspection Time | - | 0.628 | - | - |
| Four choice reaction | - | -0.773 | - | - |
| Verbal Paired Associates | - | - | - | 0.809 |
| Logical Memory | - | - | | 0.827 |
| Spatial Span | - | - | 0.739 | - |
| **Proportion of variance** | **0.498** | **0.588** | **0.628** | **0.569** |

*Note* the tests used to generate a general score of cognitive ability were used to facilitate comparison with previous work by Stevenson et al. (2019)*

**Table 5. CpG sites and relative weights** (from Lighthart et al. 2016) used to generate DNAm CRP score

| *CpG* | ***Gene*** | **Beta (discovery sample)** |
|---|---|---|
| cg06126421 | *TUBB* | -0.0052 |
| cg06690548 | *SLC7A11* | -0.0048 |
| cg10636246 | *AIM 2 & IF116* | -0.0069 |
| cg18181703 | *SOCS3* | -0.0053 |
| cg19821297 | *DNASE2* | -0.0051 |
| cg25325512 | *FGD2* | -0.0051 |
| cg27023587 | *HEATR6* | -0.005 |

**Table 6.** Sensitivity analysis: anti-inflammatory drug status, health and lifestyle factors added as covariates into inflammation-brain health regressions.

| **Brain health phenotype** | **(1) age + sex** | **(2) hypertension** | **(3) CVD history** | **(4) diabetes** | **(5) BMI** | **(6) alcoholUPW** | **(7) smoking** | **(8) Anti-inflam** |
|---|---|---|---|---|---|---|---|---|
| **TB: ICV** | **-0.197 (<0.001)** | **-0.196 (<0.001)** | **-0.194 (<0.001)** | **-0.191 (<0.001)** | **-0.183 (<0.001)** | **-0.168 (<0.001)** | **-0.167 (<0.001)** | **-0.173 (<0.001)** |
| **GM: ICV** | **-0.200 (<0.001)** | **-0.192 (<0.001)** | **-0.199 (<0.001)** | **-0.194 (<0.001)** | **-0.182 (<0.001)** | **-0.188 (<0.001)** | **-0.179 (<0.001)** | **-0.178 (<0.001)** |
| **NAWM: ICV** | **-0.150 (0.001)** | **-0.147 (0.001)** | **-0.147 (0.001)** | **-0.145 (0.002)** | **-0.138 (0.003)** | **-0.143 (0.003)** | **-0.115 (0.015)** | **-0.173 (0.004)** |
| **WMH: ICV** | **0.108 (0.017)** | **0.096 (0.026)** | **0.109 (0.014)** | **0.106 (0.019)** | **0.097 (0.029)** | **0.129 (0.005)** | 0.089 (0.05) | **0.111 (0.019)** |
| **gFA** | **-0.162 (<0.001)** | **-0.151 (0.001)** | **-0.155 (0.001)** | **-0.164 (<0.001)** | **-0.133 (0.005)** | **-0.231(<0.001)** | **-0.146 (0.004)** | **-0.149 (0.003)** |
| **gMD** | **0.124 (0.010)** | **0.121 (0.010)** | **0.123 (0.009)** | **0.124 (0.009)** | **0.141 (0.003)** | **0.172 (0.001)** | **0.114 (0.019)** | **0.109 (0.020)** |
| **visuospatial ability** | **-0.097 (0.014)** | **-0.093 (0.014)** | **-0.090 (0.015)** | **-0.089 (0.019)** | **-0.101 (0.009)** | **-0.117 (0.005)** | -0.066 (0.082) | **-0.081 (0.039)** |
| **processing speed** | **-0.144 (<0.001)** | **-0.138 (<0.001)** | **-0.139 (<0.001)** | **-0.132 (0.001)** | **-0.122 (0.003)** | **-0.179 (<0.001)** | **-0.110 (0.009)** | **-0.129 (0.002)** |
| **verbal memory** | **-0.095 (0.017)** | **-0.096 (0.014)** | **-0.099 (0.012)** | **-0.090 (0.019)** | **-0.095 (0.017)** | **-0.135 (0.003)** | -0.080 (0.050) | **-0.095 (0.020)** |
| ***gf*** | **-0.158 (<0.001)** | **-0.154(<0.001)** | **-0.154 (<0.001)** | **-0.150 (<0.001)** | **-0.148 (<0.001)** | **-0.184 (<0.001)** | **-0.129 (0.002)** | **-0.148 (<0.001)** |

*Note:* Regression models with added health covariates entered individually. Standardised betas (p values) reported. bold typeface denotes $p < 0.05$ (FDR corrected).
The following regressions are reported:
[column 1] Brain health phenotype ~ DNAm CRP + age + sex
[column 2] Brain health phenotype ~ DNAm CRP + age + sex + hypertension
[column 3] Brain health phenotype ~ DNAm CRP + age + sex + CVD history
[column 4] Brain health phenotype ~ DNAm CRP + age + sex + diabetes
[column 5] Brain health phenotype ~ DNAm CRP + age + sex + BMI
[column 6] Brain health phenotype ~ DNAm CRP + age + sex + alcohol units per week
[column 7] Brain health phenotype ~ DNAm CRP + age + sex + smoking
[column 8] Brain health phenotype ~ DNAm CRP + age + sex + anti-inflammatory drug status
Note that WMH, visuospatial ability and verbal memory are no longer significant when smoking is accounted for in the model, post FDR correction (in contrast to significant association previously).

**Table 7: FA white matter tract associations with inflammation measures**

| White matter tract | Side of hemisphere | Serum CRP (model 1) | | | Epigenetic CRP (model 2) | | | Δ association magnitudes |
|---|---|---|---|---|---|---|---|---|
| | | **β** | **SE** | **p** | **β** | **SE** | **p** | **p** |
| gFA | | -0.055 | 0.045 | 0.713 | -0.162 | 0.045 | **0.005** | **0.045** |
| Corpus callosum | Genu | -0.016 | 0.044 | 0.999 | -0.119 | 0.045 | **0.020** | **0.037** |
| | Splenium | -0.090 | 0.044 | 0.302 | -0.078 | 0.044 | 0.104 | 0.817 |
| Arcuate fasciculus | Left | 0.002 | 0.043 | 0.999 | -0.108 | 0.044 | **0.031** | **0.030** |
| | Right | -0.053 | 0.045 | 0.713 | -0.150 | 0.046 | **0.008** | 0.062 |
| Anterior thalamic radiation | Left | -0.036 | 0.045 | 0.830 | -0.120 | 0.044 | **0.020** | 0.100 |
| | Right | -0.039 | 0.043 | 0.830 | -0.126 | 0.044 | **0.015** | 0.084 |
| Rostral cingulum | Left | -0.006 | 0.044 | 0.999 | -0.050 | 0.044 | 0.317 | 0.389 |
| | Right | -0.124 | 0.045 | 0.089 | -0.140 | 0.045 | **0.009** | 0.753 |
| Uncinate fasciculus | Left | 0.001 | 0.044 | 0.999 | -0.106 | 0.045 | **0.037** | **0.040** |
| | Right | -0.034 | 0.044 | 0.830 | -0.099 | 0.044 | **0.042** | 0.197 |
| Inferior longitudinal fasciculus | Left | $-3.31 \times 10^{-5}$ | 0.043 | 0.999 | -0.032 | 0.044 | 0.499 | 0.531 |
| | Right | -0.073 | 0.044 | 0.496 | -0.084 | 0.043 | 0.081 | 0.825 |

**Table 8: MD white matter tract associations with inflammation measures**

| White matter tract | Side of hemisphere | Serum CRP (model 1) | | | Epigenetic CRP (model 2) | | | Δ association magnitudes |
|---|---|---|---|---|---|---|---|---|
| | | **β** | **SE** | **p** | **β** | **SE** | **p** | **p** |
| gMD | | -0.025 | 0.045 | 0.758 | 0.124 | 0.045 | **0.043** | **0.056** |
| Corpus callosum | Genu | -0.034 | 0.044 | 0.758 | 0.060 | 0.045 | 0.301 | 0.067 |
| | Splenium | 0.056 | 0.043 | 0.730 | 0.079 | 0.044 | 0.139 | 0.650 |
| Arcuate fasciculus | Left | -0.028 | 0.043 | 0.758 | 0.105 | 0.044 | 0.062 | **0.009** |
| | Right | 0.018 | 0.044 | 0.758 | 0.111 | 0.045 | 0.062 | 0.075 |
| Anterior thalamic radiation | Left | -0.029 | 0.045 | 0.758 | 0.083 | 0.044 | 0.139 | **0.029** |
| | Right | -0.022 | 0.043 | 0.758 | 0.016 | 0.044 | 0.817 | 0.452 |
| Rostral cingulum | Left | 0.074 | 0.044 | 0.610 | 0.078 | 0.043 | 0.139 | 0.940 |
| | Right | 0.074 | 0.044 | 0.610 | 0.093 | 0.044 | 0.099 | 0.717 |
| Uncinate fasciculus | Left | -0.015 | 0.046 | 0.758 | 0.046 | 0.046 | 0.433 | 0.243 |
| | Right | 0.037 | 0.043 | 0.758 | 0.139 | 0.044 | **0.024** | **0.045** |
| Inferior longitudinal fasciculus | Left | -0.066 | 0.043 | 0.610 | 0.054 | 0.044 | 0.329 | **0.018** |
| | Right | 0.022 | 0.045 | 0.758 | 0.013 | 0.044 | 0.817 | 0.868 |

*Note* **Supplementary Tables 7-8.** Results of linear regression analyses of epigenetic CRP score and serum CRP score with white matter tract microstructural metrics (FA and MD. Furthest right column displays difference between association magnitudes of both models, as assessed by Williams' test.

**Table 9.** Sensitivity analysis: anti-inflammatory drug status, health and lifestyle factors added as covariates into inflammation-white matter tract regressions (FA).

| White matter tract | Side | (1) age + sex | | (2) hypertension | | (3) CVD history | | (4) diabetes | | (5) BMI | | (6) alcohol UPW | | (7) smoking | | (8) anti_inflam | |
|---|---|---|---|---|---|---|---|---|---|---|---|---|---|---|---|---|---|
| | | **β** | **p** | **β** | **p** | **β** | **p** | **β** | **p** | **β** | **p** | **β** | **p** | **β** | **p** | **β** | **p** |
| Corpus callosum | Genu | -0.119 | **0.020** | -0.117 | **0.029** | -0.116 | **0.026** | -0.124 | **0.019** | -0.111 | 0.053 | -0.149 | **0.006** | -0.106 | **0.046** | -0.120 | **0.027** |
| | Splenium | -0.078 | 0.104 | -0.072 | 0.138 | -0.074 | 0.129 | -0.079 | 0.104 | -0.054 | 0.292 | -0.084 | 0.099 | -0.050 | 0.362 | -0.069 | 0.196 |
| Arcuate fasciculus | Left | -0.108 | **0.031** | -0.100 | **0.050** | -0.102 | **0.045** | -0.112 | **0.025** | -0.096 | 0.071 | -0.164 | **0.001** | -0.109 | **0.046** | -0.086 | 0.110 |
| | Right | -0.150 | **0.008** | -0.110 | **0.031** | -0.117 | **0.025** | -0.114 | **0.025** | -0.108 | 0.053 | -0.169 | **0.001** | -0.124 | **0.034** | -0.131 | **0.020** |
| Anterior thalamic radiation | Left | -0.120 | **0.020** | -0.051 | 0.308 | -0.048 | 0.347 | -0.055 | 0.271 | -0.031 | 0.560 | -0.060 | 0.255 | -0.046 | 0.389 | -0.045 | 0.407 |
| | Right | -0.126 | **0.015** | -0.037 | 0.460 | -0.036 | 0.476 | -0.040 | 0.417 | -0.055 | 0.292 | -0.090 | 0.095 | -0.041 | 0.426 | -0.038 | 0.463 |
| Rostral cingulum | Left | -0.050 | 0.317 | -0.029 | 0.550 | -0.027 | 0.579 | -0.032 | 0.500 | -0.025 | 0.609 | -0.090 | 0.095 | -0.022 | 0.664 | -0.017 | 0.753 |
| | Right | -0.140 | **0.009** | -0.101 | **0.050** | -0.101 | 0.051 | -0.103 | **0.041** | -0.091 | 0.096 | -0.166 | **0.001** | -0.097 | 0.072 | -0.109 | **0.047** |
| Uncinate fasciculus | Left | -0.106 | **0.037** | -0.134 | **0.014** | -0.142 | **0.009** | -0.143 | **0.008** | -0.128 | **0.026** | -0.188 | **0.001** | -0.120 | **0.034** | -0.133 | **0.020** |
| | Right | -0.099 | **0.042** | -0.018 | 0.687 | -0.015 | 0.740 | -0.020 | 0.647 | -0.002 | 0.958 | -0.009 | 0.857 | 0.003 | 0.943 | -0.003 | 0.954 |
| Inferior longitudinal fasciculus | Left | -0.032 | 0.499 | -0.076 | 0.118 | -0.078 | 0.112 | -0.087 | 0.071 | -0.067 | 0.196 | -0.094 | 0.094 | -0.072 | 0.165 | -0.057 | 0.276 |
| | Right | -0.084 | 0.081 | -0.094 | 0.058 | -0.091 | 0.066 | -0.103 | **0.039** | -0.077 | 0.152 | -0.119 | **0.027** | -0.088 | 0.092 | -0.080 | 0.143 |

*Note:* Inflammation-FA regression models with added health covariates entered individually. Standardised betas (p values) reported. bold typeface denotes $p < 0.05$ (FDR corrected). The following regressions are reported:
[column 1] Brain health phenotype ~ DNAm CRP + age + sex
[column 2] Brain health phenotype ~ DNAm CRP + age + sex + hypertension
[column 3] Brain health phenotype ~ DNAm CRP + age + sex + CVD history
[column 4] Brain health phenotype ~ DNAm CRP + age + sex + diabetes
[column 5] Brain health phenotype ~ DNAm CRP + age + sex + BMI
[column 6] Brain health phenotype ~ DNAm CRP + age + sex + alcohol units per week
[column 7] Brain health phenotype ~ DNAm CRP + age + sex + smoking
[column 8] Brain health phenotype ~ DNAm CRP + age + sex + anti-inflammatory drug status

**Table 10.** Sensitivity analysis: anti-inflammatory drug status, health and lifestyle factors added as covariates into inflammation-white matter tract regressions (MD).

| **White matter tract** | **Side** | **(1) age + sex** | | **(2) hypertension** | | **(3) CVD history** | | **(4) diabetes** | | **(5) BMI** | | **(6) alcohol UPW** | | **(7) smoking** | | **(8) anti_inflam** | |
|---|---|---|---|---|---|---|---|---|---|---|---|---|---|---|---|---|---|
| | | **β** | **p** | **β** | **p** | **β** | **p** | **β** | **p** | **β** | **p** | **β** | **p** | **β** | **p** | **β** | **p** |
| Corpus callosum | Genu | -0.119 | **0.020** | 0.064 | 0.257 | 0.064 | 0.262 | 0.062 | 0.268 | 0.083 | 0.114 | 0.095 | 0.103 | 0.054 | 0.402 | 0.075 | 0.235 |
| | Splenium | -0.078 | 0.104 | 0.076 | 0.165 | 0.085 | 0.139 | 0.080 | 0.158 | 0.080 | 0.114 | 0.103 | 0.093 | 0.055 | 0.402 | 0.068 | 0.263 |
| Arcuate fasciculus | Left | -0.108 | **0.031** | 0.098 | 0.092 | 0.101 | 0.074 | 0.102 | 0.079 | 0.112 | **0.049** | 0.156 | **0.006** | 0.099 | 0.133 | 0.082 | 0.218 |
| | Right | -0.150 | **0.008** | 0.080 | 0.154 | 0.082 | 0.141 | 0.077 | 0.163 | 0.107 | 0.051 | 0.102 | 0.093 | 0.089 | 0.161 | 0.088 | 0.211 |
| Anterior thalamic radiation | Left | -0.120 | **0.020** | 0.081 | 0.154 | 0.075 | 0.157 | 0.083 | 0.145 | 0.086 | 0.101 | 0.099 | 0.093 | 0.070 | 0.250 | 0.057 | 0.331 |
| | Right | -0.126 | **0.015** | -0.025 | 0.721 | -0.024 | 0.739 | -0.031 | 0.609 | 0.010 | 0.887 | 0.011 | 0.819 | -0.027 | 0.641 | -0.021 | 0.755 |
| Rostral cingulum | Left | -0.050 | 0.317 | 0.055 | 0.318 | 0.059 | 0.269 | 0.059 | 0.268 | 0.086 | 0.101 | 0.095 | 0.103 | 0.050 | 0.402 | 0.035 | 0.602 |
| | Right | -0.140 | **0.009** | 0.043 | 0.473 | 0.043 | 0.476 | 0.041 | 0.510 | 0.067 | 0.207 | 0.068 | 0.248 | 0.049 | 0.407 | 0.052 | 0.420 |
| Uncinate fasciculus | Left | -0.106 | **0.037** | 0.093 | 0.101 | 0.098 | 0.074 | 0.088 | 0.133 | 0.104 | 0.051 | 0.122 | 0.058 | 0.080 | 0.188 | 0.077 | 0.219 |
| | Right | -0.099 | **0.042** | -0.009 | 0.831 | -0.010 | 0.824 | -0.018 | 0.733 | -0.006 | 0.901 | -0.040 | 0.494 | -0.027 | 0.641 | -0.023 | 0.755 |
| Inferior longitudinal fasciculus | Left | -0.032 | 0.499 | 0.010 | 0.831 | 0.011 | 0.824 | 0.019 | 0.733 | 0.030 | 0.633 | 0.018 | 0.784 | 0.001 | 0.975 | 0.004 | 0.927 |
| | Right | -0.084 | 0.081 | 0.137 | **0.030** | 0.136 | **0.032** | 0.136 | **0.034** | 0.150 | **0.014** | 0.144 | **0.014** | 0.136 | **0.044** | 0.130 | 0.074 |

Note: Inflammation-MD regression models with added health covariates entered individually. Standardised betas (p values) reported. bold typeface denotes $p < 0.05$ (FDR corrected). The following regressions are reported:
[column 1] Brain health phenotype ~ DNAm CRP + age + sex
[column 2] Brain health phenotype ~ DNAm CRP + age + sex + hypertension
[column 3] Brain health phenotype ~ DNAm CRP + age + sex + CVD history
[column 4] Brain health phenotype ~ DNAm CRP + age + sex + diabetes
[column 5] Brain health phenotype ~ DNAm CRP + age + sex + BMI
[column 6] Brain health phenotype ~ DNAm CRP + age + sex + alcohol units per week
[column 7] Brain health phenotype ~ DNAm CRP + age + sex + smoking
[column 8] Brain health phenotype ~ DNAm CRP + age + sex + anti-inflammatory drug status

**Table 11: Bivariate associations among study variables**

| Study variables | (1) | (2) | (3) | (4) | (5) | (6) | (7) | (8) | (9) | (10) | (11) | (12) | (13) | (14) | (15) | (16) | (17) | (18) | (19) |
|---|---|---|---|---|---|---|---|---|---|---|---|---|---|---|---|---|---|---|---|
| **(1) DNAm CRP** | 1.000 | | | | | | | | | | | | | | | | | | |
| **(2) serum CRP** | 0.288 | 1.000 | | | | | | | | | | | | | | | | | |
| **(3) genetic CRP** | -0.009 | 0.307 | 1.000 | | | | | | | | | | | | | | | | |
| **(4) BMI** | 0.189 | 0.292 | 0.049 | 1.000 | | | | | | | | | | | | | | | |
| **(5) hypertension** | 0.059 | 0.047 | 0.108 | 0.208 | 1.000 | | | | | | | | | | | | | | |
| **(6) diabetes** | 0.121 | 0.027 | 0.003 | 0.162 | 0.123 | 1.000 | | | | | | | | | | | | | |
| **(7) CVD history** | 0.129 | 0.002 | 0.039 | 0.079 | 0.194 | 0.080 | 1.000 | | | | | | | | | | | | |
| **(8) smoking** | 0.221 | 0.138 | 0.019 | -0.066 | 0.023 | -0.054 | -0.028 | 1.000 | | | | | | | | | | | |
| **(9) alcohol UPW** | 0.241 | 0.060 | 0.037 | 0.014 | 0.048 | -0.013 | 0.106 | 0.047 | 1.000 | | | | | | | | | | |
| **(10) *gf*** | -0.162 | -0.092 | 0.026 | -0.080 | -0.082 | -0.088 | -0.062 | -0.153 | 0.004 | 1.000 | | | | | | | | | |
| **(11) processing speed** | -0.147 | -0.083 | 0.015 | -0.127 | -0.090 | -0.102 | -0.065 | -0.181 | 0.045 | 0.789 | 1.000 | | | | | | | | |
| **(12) visuospatial ability** | -0.072 | -0.079 | 0.058 | 0.011 | -0.074 | -0.061 | -0.056 | -0.148 | 0.057 | 0.810 | 0.546 | 1.000 | | | | | | | |
| **(13) verbal memory** | -0.116 | -0.040 | -0.003 | -0.025 | 0.005 | -0.070 | 0.006 | -0.085 | 0.013 | 0.586 | 0.382 | 0.418 | 1.000 | | | | | | |
| **(14) TB** | -0.249 | -0.026 | -0.021 | -0.117 | -0.038 | -0.104 | -0.110 | -0.164 | -0.213 | 0.172 | 0.205 | 0.093 | 0.133 | 1.000 | | | | | |
| **(15) GM** | -0.213 | -0.023 | 0.028 | -0.124 | -0.100 | -0.081 | -0.049 | -0.124 | -0.122 | 0.134 | 0.147 | 0.099 | 0.099 | 0.741 | 1.000 | | | | |
| **(16) NAWM** | -0.182 | -0.023 | -0.037 | -0.092 | -0.053 | -0.070 | -0.071 | -0.169 | -0.150 | 0.241 | 0.312 | 0.155 | 0.161 | 0.530 | 0.188 | 1.000 | | | |
| **(17) WMH** | 0.103 | 0.018 | 0.018 | 0.076 | 0.137 | 0.023 | -0.005 | 0.099 | 0.021 | -0.176 | -0.230 | -0.141 | -0.100 | 0.006 | -0.219 | -0.585 | 1.000 | | |
| **(18) gFA** | -0.160 | -0.054 | -0.043 | -0.159 | -0.146 | 0.002 | -0.086 | -0.094 | -0.108 | 0.118 | 0.159 | 0.056 | 0.060 | 0.251 | 0.210 | 0.386 | -0.370 | 1.000 | |
| **(19) gMD** | 0.126 | 0.024 | 0.034 | -0.047 | 0.049 | 0.012 | 0.019 | 0.063 | -0.009 | -0.093 | -0.148 | -0.092 | -0.124 | -0.049 | -0.105 | -0.313 | 0.428 | -0.480 | 1.000 |

*Note*. Pearson's r reported. TB: total brain volume, WMH: white matter hyperintensity volume, GM: grey matter volume; NAWM: normal appearing white matter volume. gf, general cognitive ability; gFA: general fractional anisotropy, gMD: general mean diffusivity; CVD: cardiovascular disease history

**Table 12. Results of single SEM mediation models (TB, GM, NAWM, WMH, gFA, gMD) assessing the relationship of brain structure with cognitive ability**.

| Mediator variable | Independent variable | Model | Indirect effect (ab) | | | Total effect (c) | | | Direct effect (c') | | | Attenuation |
|---|---|---|---|---|---|---|---|---|---|---|---|---|
| | | | β | SE | p | β | SE | p | β | SE | p | |
| TB volume | serum CRP | model 1 | -0.007 | 0.007 | 0.345 | -0.099 | 0.038 | **0.009** | -0.092 | 0.038 | **0.015** | 6.76% |
| | | model 2 | 0.005 | 0.008 | 0.542 | -0.066 | 0.046 | 0.150 | -0.070 | 0.045 | 0.120 | -7.25% |
| | DNAm CRP | model 1 | -0.031 | 0.011 | **0.005** | -0.158 | 0.037 | **0.000** | -0.128 | 0.038 | **0.001** | 19.46% |
| | | model 2 | -0.015 | 0.009 | 0.100 | -0.135 | 0.045 | **0.002** | -0.120 | 0.045 | **0.007** | 11.15% |
| GM volume | serum CRP | model 1 | -0.004 | 0.005 | 0.492 | -0.099 | 0.038 | **0.010** | -0.095 | 0.038 | **0.012** | 3.61% |
| | | model 2 | 0.002 | 0.005 | 0.607 | -0.065 | 0.046 | 0.153 | -0.067 | 0.046 | 0.138 | -3.67% |
| | DNAm CRP | model 1 | -0.022 | 0.010 | **0.025** | -0.158 | 0.037 | **0.000** | -0.136 | 0.038 | **0.000** | 14.02% |
| | | model 2 | -0.012 | 0.008 | 0.131 | -0.135 | 0.045 | **0.003** | -0.122 | 0.045 | **0.006** | 9.06% |
| NAWM volume | serum CRP | model 1 | -0.009 | 0.010 | 0.377 | -0.098 | 0.038 | **0.010** | -0.089 | 0.037 | **0.017** | 9.39% |
| | | model 2 | 0.003 | 0.012 | 0.777 | -0.065 | 0.046 | 0.153 | -0.069 | 0.045 | 0.125 | -5.23% |
| | DNAm CRP | model 1 | -0.033 | 0.011 | **0.003** | -0.159 | 0.037 | **0.000** | -0.125 | 0.037 | **0.001** | 21.00% |
| | | model 2 | -0.016 | 0.011 | 0.155 | -0.135 | 0.045 | **0.002** | -0.119 | 0.044 | **0.007** | 12.10% |
| WMH volume | serum CRP | model 1 | -0.004 | 0.008 | 0.653 | -0.099 | 0.038 | **0.010** | -0.095 | 0.038 | **0.011** | 3.58% |
| | | model 2 | -0.001 | 0.008 | 0.858 | -0.065 | 0.046 | 0.155 | -0.064 | 0.045 | 0.160 | 2.08% |
| | DNAm CRP | model 1 | -0.016 | 0.008 | **0.047** | -0.158 | 0.037 | **0.000** | -0.142 | 0.037 | **0.000** | 10.09% |
| | | model 2 | -0.010 | 0.007 | 0.175 | -0.134 | 0.045 | **0.003** | -0.124 | 0.045 | **0.005** | 7.55% |
| gFA volume | serum CRP | model 1 | -0.006 | 0.006 | 0.258 | -0.099 | 0.038 | **0.009** | -0.093 | 0.038 | **0.015** | 6.31% |
| | | model 2 | -0.003 | 0.006 | 0.673 | -0.066 | 0.046 | 0.150 | -0.063 | 0.045 | 0.165 | 3.86% |
| | DNAm CRP | model 1 | -0.015 | 0.008 | 0.079 | -0.158 | 0.037 | **0.000** | -0.144 | 0.038 | **0.000** | 9.19% |
| | | model 2 | -0.018 | 0.011 | 0.099 | -0.135 | 0.045 | **0.003** | -0.117 | 0.046 | **0.010** | 13.26% |
| gMD volume | serum CRP | model 1 | -0.002 | 0.004 | 0.571 | -0.098 | 0.038 | **0.010** | -0.096 | 0.038 | **0.011** | 2.10% |
| | | model 2 | -0.002 | 0.003 | 0.612 | -0.065 | 0.046 | 0.157 | -0.063 | 0.046 | 0.167 | 2.43% |
| | DNAm CRP | model 1 | -0.007 | 0.006 | 0.229 | -0.158 | 0.037 | **0.000** | -0.151 | 0.037 | **0.000** | 4.69% |
| | | model 2 | -0.006 | 0.009 | 0.518 | -0.134 | 0.045 | **0.003** | -0.128 | 0.046 | **0.005** | 4.39% |

*Note:*
Model 1 = Brain health variable ~ age + sex + inflammation;
Model 2 = Brain health variable ~ age + sex + inflammation + BMI + hypertension + smoking status + alcohol use + CVD history + diabetes

**Table 13. Results of multiple SEM mediation models (where TB, GM, NAWM, WMH, gFA, gMD are entered simultaneously) assessing the relationship of brain structure with cognitive ability**.

| Mediator variable | Independent variable | Model | Indirect effect (ab) | | | Total effect (c) | | | Direct effect (c') | | | Attenuation |
|---|---|---|---|---|---|---|---|---|---|---|---|---|
| | | | β | SE | p | β | SE | p | β | SE | p | |
| **Total [GM, NAWM, WMH, gFA, gMD]** | serum CRP | model 1 | -0.011 | 0.011 | 0.324 | -0.099 | 0.038 | **0.009** | -0.088 | 0.037 | **0.018** | 11.26% |
| | | model 2 | 0.005 | 0.013 | 0.729 | -0.066 | 0.046 | 0.150 | -0.070 | 0.045 | 0.115 | -6.96% |
| | DNAm CRP | model 1 | -0.047 | 0.015 | **0.002** | -0.159 | 0.037 | **0.000** | -0.112 | 0.038 | **0.003** | 29.39% |
| | | model 2 | -0.025 | 0.016 | 0.127 | -0.136 | 0.045 | **0.002** | -0.111 | 0.045 | **0.015** | 18.50% |
| **GM volume** | serum CRP | model 1 | -0.002 | 0.004 | 0.503 | -0.099 | 0.038 | **0.009** | -0.088 | 0.037 | **0.018** | 2.50% |
| | | model 2 | 0.002 | 0.004 | 0.635 | -0.066 | 0.046 | 0.150 | -0.070 | 0.045 | 0.115 | -2.59% |
| | DNAm CRP | model 1 | -0.015 | 0.009 | 0.097 | -0.159 | 0.037 | **0.000** | -0.112 | 0.038 | **0.003** | 9.74% |
| | | model 2 | -0.010 | 0.007 | 0.198 | -0.136 | 0.045 | **0.002** | -0.111 | 0.045 | **0.015** | 7.10% |
| **NAWM volume** | serum CRP | model 1 | -0.008 | 0.009 | 0.384 | -0.099 | 0.038 | **0.009** | -0.088 | 0.037 | **0.018** | 8.03% |
| | | model 2 | 0.002 | 0.011 | 0.824 | -0.066 | 0.046 | 0.150 | -0.070 | 0.045 | 0.115 | -3.80% |
| | DNAm CRP | model 1 | -0.029 | 0.012 | **0.012** | -0.159 | 0.037 | **0.000** | -0.112 | 0.038 | **0.003** | 18.18% |
| | | model 2 | -0.015 | 0.011 | 0.174 | -0.136 | 0.045 | **0.002** | -0.111 | 0.045 | **0.015** | 11.32% |
| **WMH volume** | serum CRP | model 1 | -0.001 | 0.002 | 0.661 | -0.099 | 0.038 | **0.009** | -0.088 | 0.037 | **0.018** | 1.08% |
| | | model 2 | 0.000 | 0.002 | 0.861 | -0.066 | 0.046 | 0.150 | -0.070 | 0.045 | 0.115 | 0.40% |
| | DNAm CRP | model 1 | -0.003 | 0.006 | 0.628 | -0.159 | 0.037 | **0.000** | -0.112 | 0.038 | **0.003** | 1.85% |
| | | model 2 | -0.001 | 0.006 | 0.900 | -0.136 | 0.045 | **0.002** | -0.111 | 0.045 | **0.015** | 0.52% |
| **gFA volume** | serum CRP | model 1 | 0.000 | 0.003 | 0.896 | -0.099 | 0.038 | **0.009** | -0.088 | 0.037 | **0.018** | 0.39% |
| | | model 2 | -0.001 | 0.002 | 0.765 | -0.066 | 0.046 | 0.150 | -0.070 | 0.045 | 0.115 | 0.94% |
| | DNAm CRP | model 1 | -0.001 | 0.008 | 0.890 | -0.159 | 0.037 | **0.000** | -0.112 | 0.038 | **0.003** | 0.73% |
| | | model 2 | -0.006 | 0.011 | 0.580 | -0.136 | 0.045 | **0.002** | -0.111 | 0.045 | **0.015** | 4.53% |
| **gMD volume** | serum CRP | model 1 | 0.000 | 0.002 | 0.806 | -0.099 | 0.038 | **0.009** | -0.088 | 0.037 | **0.018** | -0.40% |
| | | model 2 | 0.001 | 0.003 | 0.651 | -0.066 | 0.046 | 0.150 | -0.070 | 0.045 | 0.115 | -1.90% |
| | DNAm CRP | model 1 | 0.002 | 0.006 | 0.778 | -0.159 | 0.037 | **0.000** | -0.112 | 0.038 | **0.003** | -1.12% |
| | | model 2 | 0.007 | 0.010 | 0.515 | -0.136 | 0.045 | **0.002** | -0.111 | 0.045 | **0.015** | -4.97% |

*Note:*
Model 1 = Brain health variable ~ age + sex + inflammation;
Model 2 = Brain health variable ~ age + sex + inflammation + BMI + hypertension + smoking status + alcohol use + CVD history + diabetes

### Supplementary Figure 1

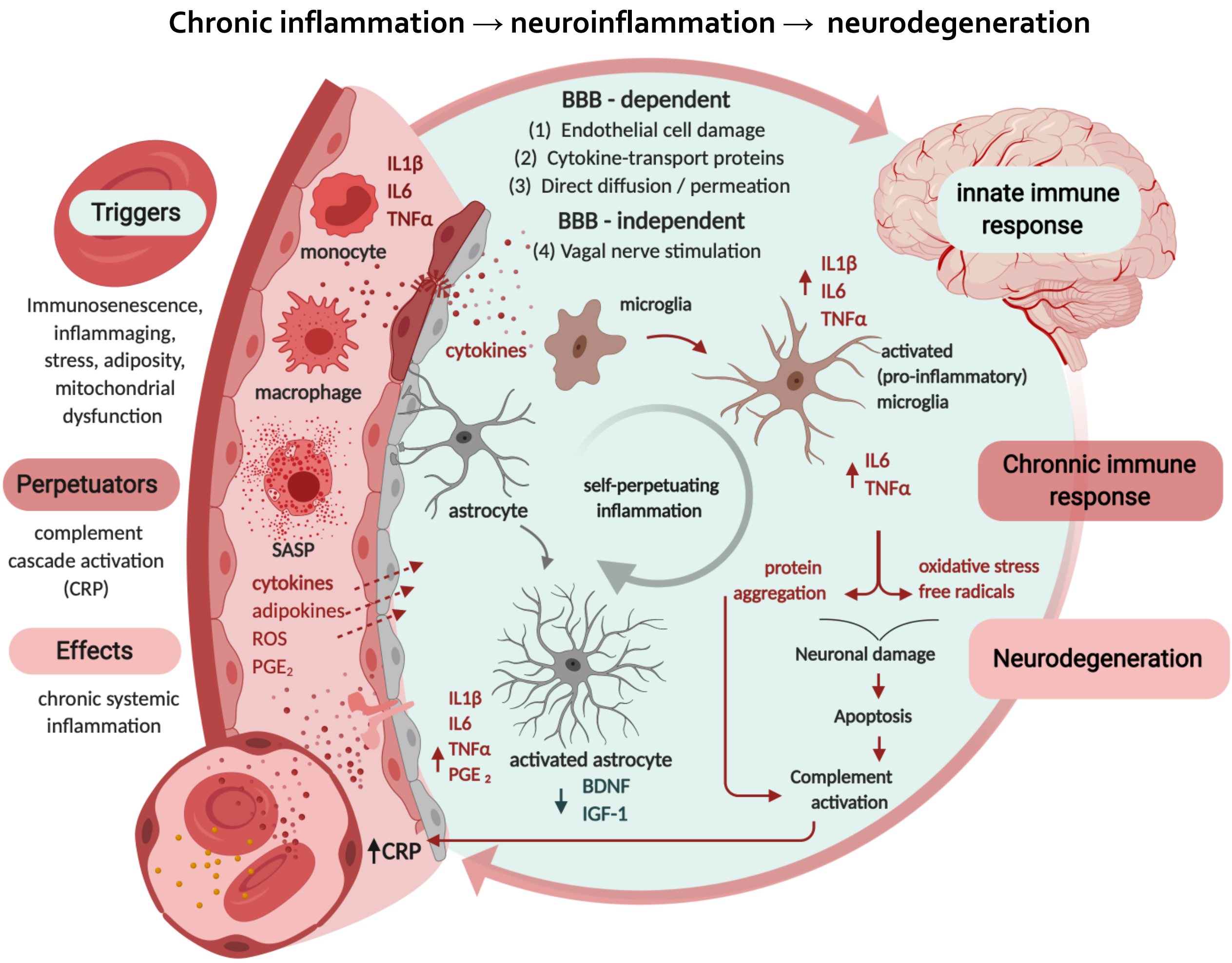


**Figure 1. Mechanisms of neurodegeneration via elevated systemic circulation.**

Increased peripheral inflammation is pertinent to brain ageing in that inflammatory mediators in the periphery can damage the blood brain barrier (BBB), permitting entry into the brain where they go on to disrupt neurons and glia and promote neuroinflammation. This directly contributes to various neurodegenerative pathways which lead to brain cell death, as outlined previously [1,2]. *BBB: Blood-brain barrier; CRP: C-reactive protein; IL6: interleukin-6; IL1 β: interleukin-1β; TNF-α: tumour necrosis factor-α; PGE₂: prostaglandin E2; BDNF: Brain-derived neurotrophic factor; IGF-1: insulin-like growth factor 1; SASP: senescence-associated secretory phenotype; ROS: reactive oxygen species*

### Supplementary Figure 2

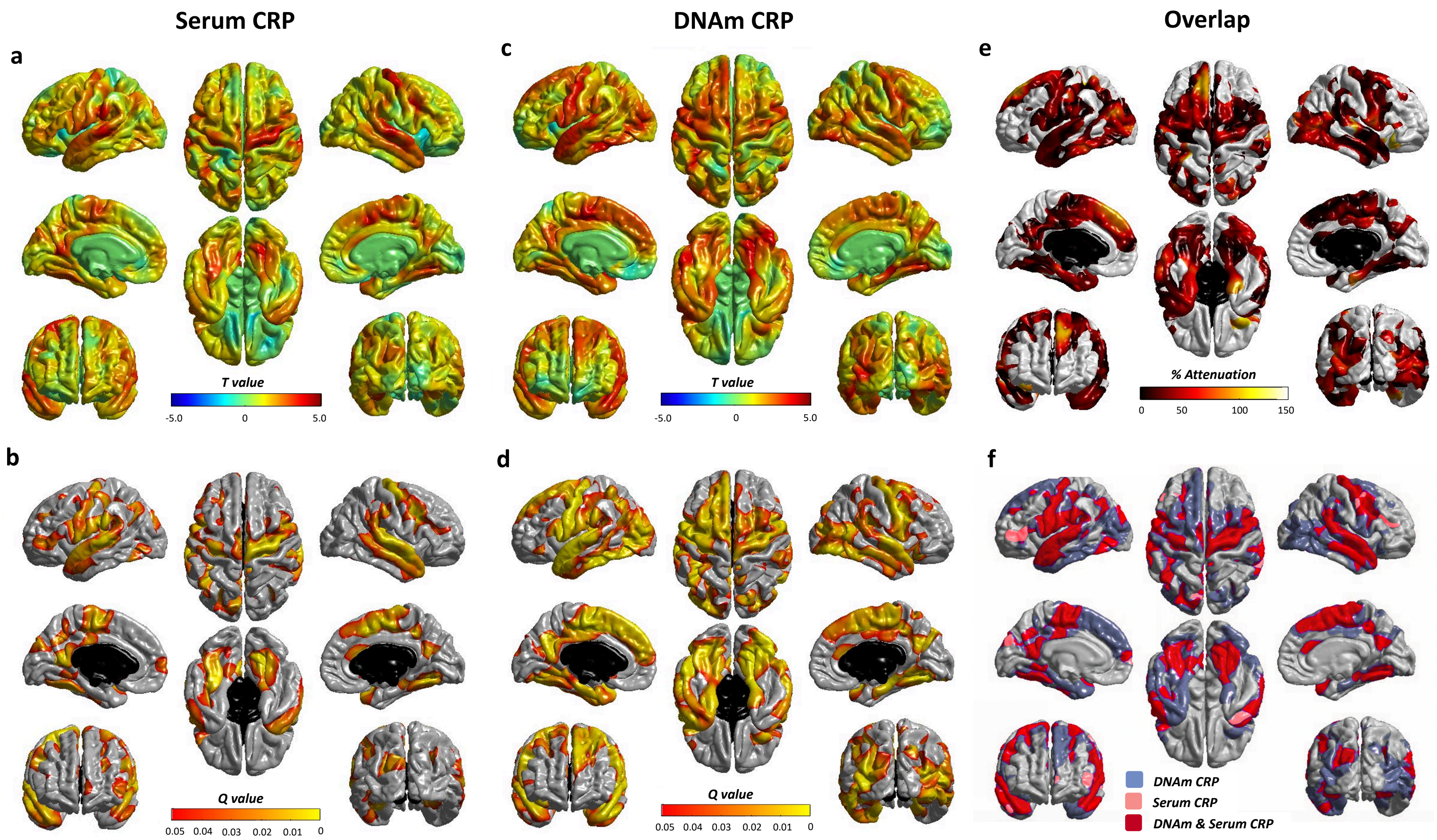


**Fig. S2. DNAm CRP shows stronger and more widespread associations with regional brain cortical thickness than serum CRP.**

Regional cortical thickness regressed against serum CRP (**i-ii**) and DNAm CRP (**iii-iv**). Colours denote the magnitude (T-maps; top) and significance (Q values; bottom) of the negative associations between inflammation and brain cortical thickness. Panel (**v**) shows the percentage attenuation for the significant associations between DNAm-CRP and cortical thickness when also controlling for serum CRP. Conjunction plot (**vi**) shows the spatial extent of independent contributions and overlap (red) in cortical loci that exhibit FDR-corrected unique associations with simultaneously-modelled serum (pink) and epigenetic (blue) inflammation measures; results are corrected for sex, age and ICV. TB: total brain, GM: grey matter, NAWM: normal-appearing white matter, WMH: white matter hyperintensity, $g_f$, general cognitive ability; gFA: general fractional anisotropy, gMD: general mean diffusivity; n=521
